# Palliative Care among People with Advanced Heart Failure: Between Hospital Variation in the US Veterans Administration

**DOI:** 10.64898/2026.07.22.26358645

**Authors:** Shelli L. Feder, Na Ouyang, Ling Han, Erica A. Abel, Eric E. DeRycke, Daniel Kinder, Nancy S. Redeker, Leslie Curry, Jennifer Ibarra, Cari R. Levy, Carol Lurhs, Dio Kavalieratos, Kathleen M. Akgün, Larry Allen

## Abstract

**Background:** Palliative care is recommended by clinical practice guidelines for patients with advanced heart failure (aHF), yet specialty palliative care (SPC) remains substantially underutilized in this population. We sought to quantify between-facility variation in SPC receipt among people with aHF and determine how much variation is explained by patient case mix and facility structural characteristics versus residual unmeasured factors.

**Methods:** This retrospective cohort study included 23,991 Veterans with prevalent aHF identified through administrative data across 133 VA Medical Centers (VAMCs) with ≥20 aHF cases, from January 2022 to December 2023. Variation was assessed using multilevel logistic regression with facility random intercepts, the intraclass correlation coefficient (ICC), and the adjusted median odds ratio (aMOR). Facility-specific risk-standardized SPC rates were used to estimate SPC encounters attributable to facility performance better or worse than the national rate.

**Results:** Of the sample, the mean patient age was 72.3 years (SD = 10.0), and 97.6% were male. The national observed rate of SPC was 17.5%, with risk-adjusted rates varying approximately 14-fold across facilities (3.3% to 45.6%). The adjusted ICC was 11.5%, and aMOR was 1.87 (95% Confidence Intervals 1.70-2.06). Measured patient case-mix and facility structural characteristics explained only 18.9% of between-facility variation (proportional reduction in the ICC, fully adjusted vs. null model). Facilities performing better than the national rate delivered 791 more SPC encounters than expected (17.8%), while those performing worse than the national rate delivered 480 fewer encounters than expected (10.8%).

**Conclusions:** In the context of a national mean rate of SPC that reflects substantial underuse, delivery varied 14-fold across VAMCs, with most variation unexplained by patient complexity or measured facility resources. These findings suggest that potentially modifiable organizational factors, beyond patient preferences or facility structures alone, may contribute to current utilization gaps and represent actionable targets for quality improvement.

**Clinical Perspective:** *What is new?:* In this national cohort of 23,991 people with advanced heart failure cared for at 133 VA Medical Centers, risk-adjusted rates of specialist palliative care delivery varied 14-fold across facilities (median odds ratio 1.87). Measured patient case-mix and facility structural characteristics explained only 18.9% of the between-facility variation.

*Clinical implications?:* Substantial variation in palliative care delivery across VAMCs is not accounted for by differences in patient complexity or facility resources, suggesting that institutional processes and referral practices are key drivers and represent actionable targets for quality improvement.

## Introduction

Heart failure affects over 6 million Americans and carries a high symptom burden, frequent hospitalization, and a five-year mortality exceeding 50%.^1–3^ Many patients progress into severe disease— advanced heart failure (aHF), stage D, end-stage, where symptoms worsen, and mortality is particularly high. Despite strong guideline recommendations, specialist palliative care (SPC), an intervention that reduces symptom burden, improves goal-concordant care, and is associated with better quality of life, remains underutilized in this population.^4–7^ Prior studies have documented variation in SPC receipt across hospitals and health systems, yet have not quantified the share of this variation potentially attributable to modifiable factors.^8,9^

The Veterans Health Administration (VA), the largest integrated healthcare system in the United States, provides care to more than 87,000 people with aHF each year. Beginning in 2003, the VA prioritized palliative care across the delivery system, and most VA Medical Centers (VAMCs) now offer these services.^10^ VA palliative care rates for aHF exceed those in non-VA settings yet remain substantially underutilized.^7^ This gap suggests that system-level factors, not only patient characteristics, may drive palliative care receipt and that improvement is achievable through targeted intervention. Barriers to palliative care referral fall into two broad domains: structural factors (e.g., palliative care team capacity, facility size, and geography) that may be difficult to change in the short term, and process and behavioral factors (e.g., clinician referral practices, prognostic communication, and care culture) that may be more tractable targets for quality improvement. Disentangling the relative contributions of these domains requires formal risk standardization that separates patient case mix from facility-level performance. Without such standardization, low utilization and apparent variation in SPC rates may simply reflect differences in patient complexity rather than differences in facility quality.

Thus, the purpose of this study was to estimate national rates of SPC among people with aHF treated in the VA. Further, we sought to quantify between-facility variation in SPC receipt, determine the extent to which variation is explained by patient case mix and facility structural characteristics versus residual unmeasured factors, and estimate the number of palliative care encounters attributable to above-and below-average facility performance. Together, these analyses identify how much variation in SPC is potentially modifiable and where improvement efforts may be most warranted.

## Materials and Methods

### Study Design

We conducted a retrospective observational study of data from the HEART-PAL cohort, a VA-based cohort designed to examine care near the end of life among people with heart failure.^7,8^ Data from HEART-PAL are collected from the electronic health record of the VA’s Corporate Data Warehouse (CDW) and the Centers for Medicare and Medicaid Services. The Institutional Review Board at the VA Connecticut Healthcare System approved this study. We followed the Reporting of Studies Conducted Using Observational Routinely Collected Data (RECORD) reporting guideline, an extension of the Strengthening the Reporting of Observational Studies in Epidemiology guidelines.^11^

### Data Sources and Study Population

Patients were identified as having aHF, using validated administrative algorithms, through a lookback period spanning 2017–2022.^12^ Specifically, the algorithm used two hospitalizations (admitted >24 hours) with a discharge diagnosis of heart failure, ventricular arrhythmia, or advanced therapies (e.g., left ventricular assist device) by International Classification of Diseases 10^th^ Edition (ICD-10) codes and one sign of aHF defined as hyponatremia, hypotension, acute kidney injury or dialysis by ICD-10 code or Current Procedural Terminology codes or high-dose loop diuretic or metolazone use from pharmacy claims within a 12-month timeframe (Supplement). This algorithm has good performance in EHR review, with a sensitivity of 73%, specificity of 90%, PPV of 61%, and an NPV of 94%.^12^ Outcomes were assessed over a fixed two-year observation period, January 1, 2022, through December 31, 2023, for all patients, providing a uniform window in which each facility had equal opportunity to deliver palliative care. People with aHF were assigned to the VAMC, where they received the majority of their care during the study timeframe. Over 75% received care at one VAMC during the study period. The analytic sample was further restricted to facilities with at least 20 patients to ensure stable estimates.

### Outcomes and Patient and Facility-Level Covariates

We used a validated EHR-based algorithm comprising Current Procedural Terminology codes and VA-specific stop codes to identify SPC encounters during the study period (Supplement).^13^ Algorithm sensitivity is 90%, specificity 100%, PPV 100%, and NPV 93% for VA-based SPC.^13^ To capture the full array of palliative delivery, we constructed two binary outcomes: (1) receipt of any SPC (i.e., follow-up and new consultations) and (2) receipt of a new SPC consultation (i.e., no follow-up encounters included), both assessed during the 2022–2023 observation window. Covariates were collected from the 12 months before the study observation window. Patient-level covariates included age, sex, marital status, dual Medicare-Medicaid eligibility, VA service-connected disability status and VA enrollment priority group (a categorical variable based on service-connection and other eligibility criteria that determines copayment requirements and access to specific benefits), Elixhauser Comorbidity Index (calculated from ICD-10 codes from inpatient and outpatient visits up to 12 months before cohort entry), number of unique chronic medications, total emergency department visits in the prior year, the JEN Frailty Index (JFI), and the VA Care, Assessment, Needs (CAN) risk score.^14,15^ The JFI was categorized using established thresholds reflecting low, moderate, and high long-term institutionalization risk, and CAN scores were categorized using established VA operations cut-points at the 60th and 90th percentiles to denote elevated and high one-year mortality risk, respectively.

Cohort entry year was included as a fixed covariate to account for secular trends across the 2017– 2022 identification window. Facility-level structural covariates included a three-level VA complexity designation (Tier 1: high complexity [levels 1a–1c]; Tier 2: mid complexity [level 2]; Tier 3: low complexity [level 3]) and log-transformed facility volume. Complexity is a VA-assigned composite measure of resource availability, including bed availability, intensive care unit capacity, specialty care, and research.^16^ Higher complexity tiers reflect greater facility resources, larger patient volumes, and a broader scope of clinical and academic services. Facility panel size, defined as the log-transformed total number of unique patients with aHF receiving care at the VAMC during the study period, was included to account for differences in facility scale.

### Statistical Analysis

Patient outcomes were modeled using generalized linear mixed models with a facility random intercept to account for clustering within facilities. Three sequential models were estimated for each outcome: a null model including only a grand intercept and facility random effect, to establish baseline variance; a second model which added patient-level covariates (age, sex, marital status, dual eligibility, VA priority, Elixhauser Comorbidity Index, number of medications, prior ED visits, frailty index, mortality risk category, and cohort entry year and a third model which added the two facility structural covariates (complexity tier and log panel size). Multicollinearity among covariates was assessed using variance inflation factors prior to model fitting; all values were below conventional thresholds for concern. We calculated intraclass correlation coefficients (ICC) at each model stage to quantify the proportion of outcome variation attributable to VAMCs. The proportion of between-facility variation explained by measured covariates was calculated as the proportional reduction in the ICC from the null model to the fully adjusted model. To facilitate interpretation of the degree of variation between facilities, we also calculated the adjusted median odds ratio (aMOR). The aMOR can be interpreted as the median odds of a patient receiving SPC at one randomly chosen VAMC relative to receiving care at another, holding patient demographic and clinical features constant.

Facility performance was summarized using risk-standardized outcome rates (RSRs), following the methods of Keenan et al.^17^ Each facility’s RSR was calculated as the sum of patient-level predicted probabilities to expected events multiplied by the national unweighted facility mean. Expected events were computed as the sum of predicted probabilities with the facility random effect set to zero (i.e., the mean of the random effect distribution), reflecting the expected outcome under average facility performance given the facility’s patient case mix. Model discrimination was assessed using the C-statistic (area under the receiver operating characteristic curve). Calibration of model predictions was assessed using decile calibration plots and facility-level expected-to-observed ratios stratified by facility complexity.

Confidence intervals (CI) for facility RSRs for SPC use among those with aHF were estimated via a hierarchical bootstrap (1,000 iterations) in which facilities were resampled with replacement, followed by within-facility patient resampling. Percentile-based 95% CI were derived from the 2.5th and 97.5th percentiles of the 1,000 bootstrap RSR estimates for each facility. Facilities were classified as better than the national rate, worse than the national rate, or no different than the national rate based on whether the 95% bootstrap CI for the RSR fell entirely above, entirely below, or crossed the national mean. Funnel plots of RSRs were constructed with 95% and 99% control limits derived from the bootstrap distribution, plotted against facility panel size.

Attributable risk was defined as the excess palliative care events at VAMCs performing better than the national rate relative to the reference (i.e., the number of encounters occurring because certain facilities performed better than the national rate) and the counterfactual shortfall at facilities performing worse than the national rate (i.e., encounters that did not occur because certain facilities performed below the national mean of SPC for aHF). Attributable risk was computed by comparing each patient’s predicted probability under the full model (including facility random effect) to their predicted probability at a reference facility with the random effect set to zero (the population mean). Positive and negative excess events were summed within and across facilities and expressed as percentages of total observed events.

To evaluate the robustness of the primary findings, we conducted two sensitivity analyses. First, we estimated RSRs using patient-level covariates only to assess whether adding facility-level structural covariates materially changed facility classifications. Second, we replaced the national mean reference with complexity-tier-specific means, comparing each facility against the average of its structural peers rather than the national average. Because the medium- and low-complexity tiers contained only 23 and 26 facilities, respectively, we assessed the stability of the tier-specific reference using a leave-one-out analysis in which the largest facility in each tier was excluded, and the peer means were recomputed. RSR correlations and classification agreement rates were computed across all model pairs. All analyses were conducted in SAS 9.4 (SAS Institute, Cary, NC).

## Results

### Sample Characteristics

The sample included 23,991 people with aHF who received care at 133 national VAMCs (Table 1). The sample’s mean age was 73.9 years (SD = 9.8). 97.6% were male, 69.8% were White, 29.8% were Black, and 4.8% were Hispanic. Of the sample, 48% were married, 12.7% were dual-eligible, 48.6% were rated as having high disability, and 69.6% had CAN scores of 90 or higher, indicating individuals were in the 90^th^ percentile of risk for hospitalization or death within the next year. Mean Elixhauser scores were 7.5 (SD = 3.2); the number of medications was 9.2 (SD = 4.2); ED visits were 4.6 (SD = 4.0); and the JEN Frailty Index was 6.4 (SD = 2.3). Facility volume ranged from 23 to 647 patients with aHF during the study period (median 165; IQR 81–243). Facilities were distributed across complexity tiers: 84 high-complexity (VA 1a/1b/1c), 23 medium-complexity, and 26 low-complexity.

**Table 1:** Patient Characteristics by Palliative Care Outcomes

| Variables | Total Sample |  | Any Palliative Care Encounters |  |  | New Palliative Care Encounters |  |  |
| --- | --- | --- | --- | --- | --- | --- | --- | --- |
|  |  |  | No Palliative Care<br>(N=19556) | Any Palliative Care<br>(N=4435) |  | No Consult<br>(N=19801) | New Consult<br>(N=4190) |  |
|  | N | Mean (SD) or % | Mean (SD) or N (%) | Mean (SD) or N (%) | P value | Mean (SD) or N (%) | Mean (SD) or N (%) | P value |
| Age (Years) | 23991 | 73.9 (9.8) | 73.7 (9.8) | 74.5 (10.0) | <.001 | 73.8 (9.8) | 74.4 (10.1) | <.001 |
| Male | 23426 | 97.6% | 19112 (97.7%) | 4314 (97.3%) | .071 | 19350 (97.7%) | 4076 (97.3%) | .085 |
| Race (proportion, %) |  |  |  |  | .025 |  |  | .003 |
| White | 14837 | 61.8% | 12158 (62.2%) | 2679 (60.4%) |  | 12313 (62.2%) | 2524 (60.2%) |  |
| Black | 7152 | 29.8% | 5737 (29.3%) | 1415 (31.9%) |  | 5798 (29.3%) | 1354 (32.3%) |  |
| American Indian or Alaska Native | 151 | 0.6% | 126 (0.6%) | 25 (0.6%) |  | 129 (0.7%) | 22 (0.5%) |  |
| Asian | 91 | 0.4% | 76 (0.4%) | 15 (0.3%) |  | 76 (0.4%) | 15 (0.4%) |  |
| Native Hawaiian or Other Pacific Islander | 195 | 0.8% | 166 (0.9%) | 29 (0.7%) |  | 167 (0.8%) | 28 (0.7%) |  |
| Mixed | 216 | 0.9% | 184 (0.9%) | 32 (0.7%) |  | 188 (1.0%) | 28 (0.7%) |  |
| Unknown | 1349 | 5.6% | 1109 (5.7%) | 240 (5.4%) |  | 1130 (5.7%) | 219 (5.2%) |  |
| Hispanic | 1144 | 4.8% | 850 (4.3%) | 294 (6.6%) | <.001 | 870 (4.4%) | 274 (6.5%) | <.001 |
| Marital status (proportion, %) |  |  |  |  | <0.001 |  |  | <0.001 |
| Married | 11714 | 48.8% | 9683 (49.5%) | 2005 (45.2%) |  | 9814 (49.6%) | 1900 (45.4%) |  |
| Divorced/Separated/Widowed | 2504 | 10.4% | 2031 (10.4%) | 499 (11.3%) |  | 2041 (10.3%) | 463 (11.1%) |  |
| Single | 9773 | 40.7% | 7868 (40.2%) | 1905 (43.0%) |  | 7946 (40.1%) | 1827 (43.6%) |  |
| Medicare (proportion, %) |  |  |  |  | .028 |  |  | .031 |
| No Medicare | 1258 | 7.9% | 1002 (5.1%) | 256 (5.8%) |  | 1010 (5.1%) | 248 (5.9%) |  |
| Medicare only | 19053 | 79.4% | 15507 (79.3%) | 3546 (80.0%) |  | 15717 (79.4%) | 3336 (79.6%) |  |
| Dual Eligible | 3680 | 12.7% | 3047 (15.6%) | 633 (14.3%) |  | 3074 (15.5%) | 606 (14.5%) |  |
| Disability (proportion, %) |  |  |  |  | <.001 |  |  | <.001 |
| High | 11652 | 48.6% | 9375 (47.9%) | 2277 (51.3%) |  | 9511 (48.0%) | 2141 (51.1%) |  |
| Low-moderate | 3462 | 14.4% | 2856 (14.6%) | 606 (13.7%) |  | 2884 (14.6%) | 578 (13.8%) |  |
| Low income | 5894 | 24.6% | 4747 (24.3%) | 1147 (25.9%) |  | 4798 (24.2%) | 1096 (26.2%) |  |
| No service connection | 2745 | 11.4% | 2393 (12.2%) | 352 (7.9%) |  | 2421 (12.2%) | 423 (10.1%) |  |
| Unknown/Missing | 238 | 1.0% | 185 (1.0%) | 53 (1.2%) |  | 187 (0.9%) | 51 (1.2%) |  |
| Elixhauser Index | 23991 | 7.5 (3.2) | 7.4 (3.1) | 8.3 (3.1) | <.001 | 7.4 (3.1) | 8.4 (3.2) | <.001 |
| Number of medications | 23991 | 9.2 (4.2) | 8.8 (4.2) | 10.6 (3.9) | <.001 | 8.9 (4.2) | 10.6 (3.9) | <.001 |
| ED visits in the past year | 23991 | 4.6 (4.0) | 4.4 (3.8) | 5.2 (4.7) | <.001 | 4.4 (3.8) | 5.2 (4.8) | <.001 |
| JEN Frailty Index | 23991 | 6.4 (2.3) | 6.2 (2.2) | 7.1 (2.1) | <.001 | 6.2 (2.2) | 7.0 (2.1) | <.001 |
| ICU admissions total past year | 23991 | 0.6 (1.1) | 0.6 (1.1) | 0.6 (1.1) | .628 | 0.6 (1.1) | 0.6 (1.1) | .612 |
| Hospital Admissions in the Past Year | 23991 | 3.7 (2.2) | 3.6 (2.2) | 4.1 (2.4) | <.001 | 3.6 (2.1) | 4.1 (2.4) | <.001 |
| 1-Year Mortality Risk Category (CAN score)<br>(proportion, %) |  |  |  |  | <.001 |  |  | <.001 |
| CAN score <=60 | 306 | 1.3% | 283 (1.5%) | 23 (0.5%) |  | 283 (1.4%) | 23 (0.6%) |  |
| 60< CAN score <=90 | 6321 | 26.4% | 5801 (29.7%) | 520 (11.7%) |  | 5830 (29.4%) | 491 (11.7%) |  |
| CAN score >90 | 16995 | 70.8% | 13183 (67.4%) | 3812 (86.0%) |  | 13397 (67.7%) | 3598 (85.9%) |  |
| Unknown/Missing | 369 | 1.5% | 289 (1.5%) | 80 (1.8%) |  | 291 (1.5%) | 78 (1.9%) | <.001 |
| Index Year (proportion, %) |  |  |  |  | <.001 |  |  |  |
| 2017 | 2262 | 9.4% | 1787 (9.1%) | 475 (10.7%) |  | 1798 (9.1%) | 464 (11.1%) |  |
| 2018 | 3152 | 13.1% | 2584 (13.2%) | 568 (12.8%) |  | 2607 (13.2%) | 545 (13.0%) |  |
| 2019 | 4141 | 17.3% | 3376 (17.3%) | 765 (17.3%) |  | 3410 (17.2%) | 731 (17.5%) |  |
| 2020 | 3980 | 16.6% | 3235 (16.5%) | 745 (16.8%) |  | 3274 (16.5%) | 706 (16.9%) |  |
| 2021 | 5539 | 23.1% | 4485 (22.9%) | 1054 (23.8%) |  | 4559 (23.0%) | 980 (23.4%) |  |
| 2022 | 4917 | 20.5% | 4089 (20.9%) | 828 (18.7%) |  | 4153 (21.0%) | 764 (18.2%) |  |
| Facility Complexity |  |  |  |  | <.001 |  |  | <.001 |
| 1 (1a, 1b, 1c) | 20654 | 86.1% | 16679 (85.3%) | 3975 (89.6%) |  | 16885 (85.3%) | 3769 (90.0%) |  |
| 2 | 2047 | 8.5% | 1692 (8.7%) | 355 (8.0%) |  | 1725 (8.7%) | 322 (7.7%) |  |
| 3 | 1290 | 5.4% | 1185 (6.1%) | 105 (2.4%) |  | 1191 (6.0%) | 99 (2.4%) |  |
| Census Region |  |  |  |  | <.001 |  |  | <.001 |
| South | 11046 | 46.0% | 8974 (45.9%) | 2072 (46.7%) |  | 9082 (45.9%) | 1964 (46.9%) |  |
| Southeast | 5419 | 22.6% | 4454 (22.8%) | 965 (21.8%) |  | 4516 (22.8%) | 903 (21.6%) |  |
| Northeast | 3169 | 13.2% | 2564 (13.1%) | 605 (13.6%) |  | 2606 (13.2%) | 563 (13.4%) |  |
| West | 4159 | 17.3% | 3459 (17.7%) | 700 (15.8%) |  | 3492 (17.6%) | 667 (15.9%) |  |
| Other | 198 | 0.8% | 105 (0.5%) | 93 (2.1%) |  | 105 (0.5%) | 93 (2.2%) |  |
| Urban/Rural/Highly Rural |  |  |  |  | <.001 |  |  | <.001 |
| Missing | 6 | 0.0% | 6 (0.0%) | 0 (0.0%) |  | 6 (0.03%) | 0 (0.0%) |  |
| Highly Rural | 178 | 0.7% | 140 (0.7%) | 38 (0.9%) |  | 141 (0.7%) | 37 (0.9%) |  |
| Isolated | 5 | 0.0% | 5 (0.0%) | 0 (0.0%) |  | 5 (0.0%) | 0 (0.0%) |  |
| Rural | 6617 | 27.6% | 5544 (28.4%) | 1073 (24.2%) |  | 5618 (28.4%) | 999 (23.8%) |  |
| Urban | 17186 | 71.6% | 13861 (70.9%) | 3324 (75.0%) |  | 14031 (70.9%) | 3154 (75.3%) |  |
| Receipt of any palliative care | 4435 | 18.5% | -- | -- | -- | -- | -- | -- |
| New palliative care consultation | 4190 | 17.5% | -- | -- | -- | -- | -- | -- |

Of the 23,991 patients, 4,435 (18.5%) had an SPC encounter during the study period. Patients who received any SPC differed from those who did not on several baseline characteristics. Those receiving SPC were significantly older (74.5 vs. 73.7 years, p<.001). Racial composition differed significantly (p=.025), with Black patients comprising a larger proportion of SPC recipients (31.9% vs. 29.3%), as were Hispanic patients (6.6% vs. 4.3%, p<.001). Medicare status differed slightly (p=.028), with marginally more Medicare-only patients among recipients (80.0% vs. 79.3%). Disability category also differed (p<.001), with a higher proportion of high-disability patients receiving SPC (51.3% vs. 47.9%) and fewer with no service connection (7.9% vs. 12.2%).

Patients receiving SPC had greater clinical complexity, including higher Elixhauser scores (8.3 vs. 7.4, p<.001), higher JEN Frailty Index scores (7.1 vs. 6.2, p<.001), more medications (10.6 vs. 8.8, p<.001), and were more likely to have a CAN score >90 (86.0% vs. 67.4%, p<.001), more ED visits (5.2 vs. 4.4, p<.001), and more hospital admissions (4.1 vs. 3.6, p<.001). ICU admissions did not differ significantly (0.6 vs. 0.6, p=.628).

Patients were more likely to be cared for at high complexity facilities (89.6% vs. 85.3%, p<.001) and in urban settings (75.0% vs. 70.9%, p<.001). Regional distribution differed significantly (p<.001), with slightly more recipients in the South (46.7% vs. 45.9%) and fewer in the West (15.8% vs. 17.7%). Similar patterns were observed for new SPC consults.

### Crude and Risk-Adjusted Rates of Palliative Care

The national observed rate of any SPC was 18.5% (facility range, 0–47%), and 17.5% for new SPC encounters (facility range, 0–47%). In the fully adjusted multilevel model, facility-level risk-standardized rates varied substantially: from 3.3% (95% CI, 3.1–3.6%) to 45.6% (95% CI, 44–47.3%) for any SPC, and from 3.3% (95% CI, 3.1–3.6%) to 45.5% (95% CI, 43.9–47.1%) for new SPC encounters — an approximately 14-fold difference between the lowest- and highest-performing facilities for both outcomes, respectively (Figure 1, Panel A). Variation was evident within all facility complexity tiers (Figure 1, Panel B), with high-, medium-, and low-complexity facilities spanning ranges of 3.3%-45.6%, 4.2%-37%, and 5.3%-26.1%, respectively. Similar patterns were observed for new SPC encounters (Figure 1, Panels C & D).

**Figure 1.**
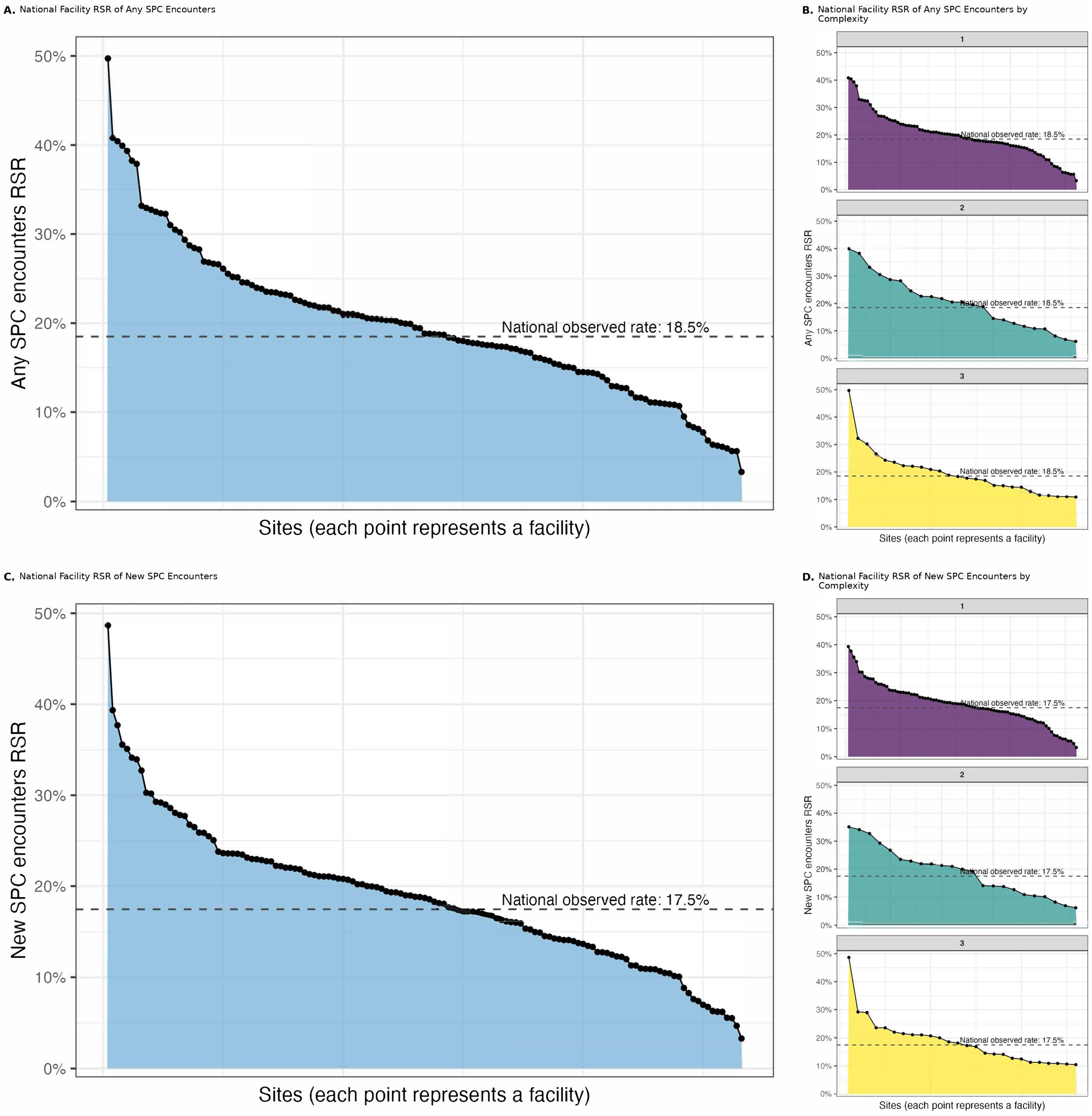
Waterfall Plots of RSR for Any and New SPC Encounters by Facility and Complexity. RSR: Risk-standardized facility rates for any Specialist Palliative Care (SPC) encounter (Panels A-B) and new SPC encounters (Panels C-D) among Veterans with advanced heart failure, 2022-2023. Panel A shows all 133 facilities ranked by performance; the horizontal dashed line indicates the national observed rate. Panels B and D stratify facilities by complexity tier. Each point represents one facility. Rates are adjusted for patient case mix and facility structural factors.

### Between-Facility Variation

In the null model, 13.8% of the total variation in any SPC was attributable to between-facility differences (τ² = 0.5267, ICC = 0.138) and 13.6% for new SPC encounters (τ² = 0.5168, ICC = 0.136). In fully adjusted models, facility-level random intercept variance was 0.4270 (ICC = 11.5%; aMOR = 1.87, 95% CI: 1.70–2.06) for any SPC and 0.4092 (ICC = 11.1%; aMOR = 1.84; 95% CI: 1.68–2.04) for new SPC encounters, respectively. Measured patient case-mix and facility structural characteristics explained 18.9% of the between-facility variation of any SPC and 20.8% of new SPC. The fully adjusted model demonstrated good discrimination (Any SPC C-statistic = 0.74; New SPC C-statistic = 0.74), and decile calibration plots and E/O ratios showed good calibration across the full range of predicted probabilities for both outcomes (E/O ratios for both outcomes of 0.93, 0.90, and 0.95 across high-, mid-, and low-complexity facilities, respectively).

### Patient and Facility Predictors of Palliative Care

In the fully adjusted model, Elixhauser (OR=1.02, 95% CI: 1.01–1.04), number of medications (OR=1.07, 95% CI: 1.06–1.08), ED visits (OR=1.02, 95% CI: 1.01–1.02), and the JEN Frailty Index (OR=1.10, 95% CI: 1.07–1.12) were associated with any SPC. CAN scores >90 had more than twice the odds of SPC receipt compared to CAN scores of ≤60 (OR=2.41, 95% CI: 1.54–3.79). Conversely, patients with Medicare-only coverage (OR=0.72, 95% CI: 0.61–0.85) or dual eligibility (OR=0.60, 95% CI: 0.50–0.72) had significantly lower odds of receiving any SPC compared to those without Medicare, as did patients with no service connection relative to high-disability patients (OR=0.80, 95% CI: 0.70–0.91). Care at high (OR=3.11, 95% CI: 1.74–5.56) and medium complexity VAMCs (OR=2.24, 95% CI: 1.38– 3.62) was associated with substantially higher odds of SPC receipt compared to low complexity VAMCs. Utilization increased significantly over time (OR=1.43, 95% CI: 1.27–1.60 per index year). Results for new SPC were similar in direction and magnitude across covariates.

### Risk-Standardized Outcome Performance

For any SPC, 63 VAMCs (47.4%) performed significantly better than the national mean, 20 (15%) performed as expected, and 50 (37.6%) performed significantly worse based on bootstrapped 95% confidence intervals (Figure 2, Panel A; Supplement, Table 2). For new SPC encounters, 67 facilities (50.4%) performed better than expected, 14 (10.5%) as expected, and 52 (39.1%) worse than expected (Figure 2. Panel B., Supplement Table 3). Poorer facility performance was distributed across levels of complexity for both outcomes (P>.05).

**Figure 2.**
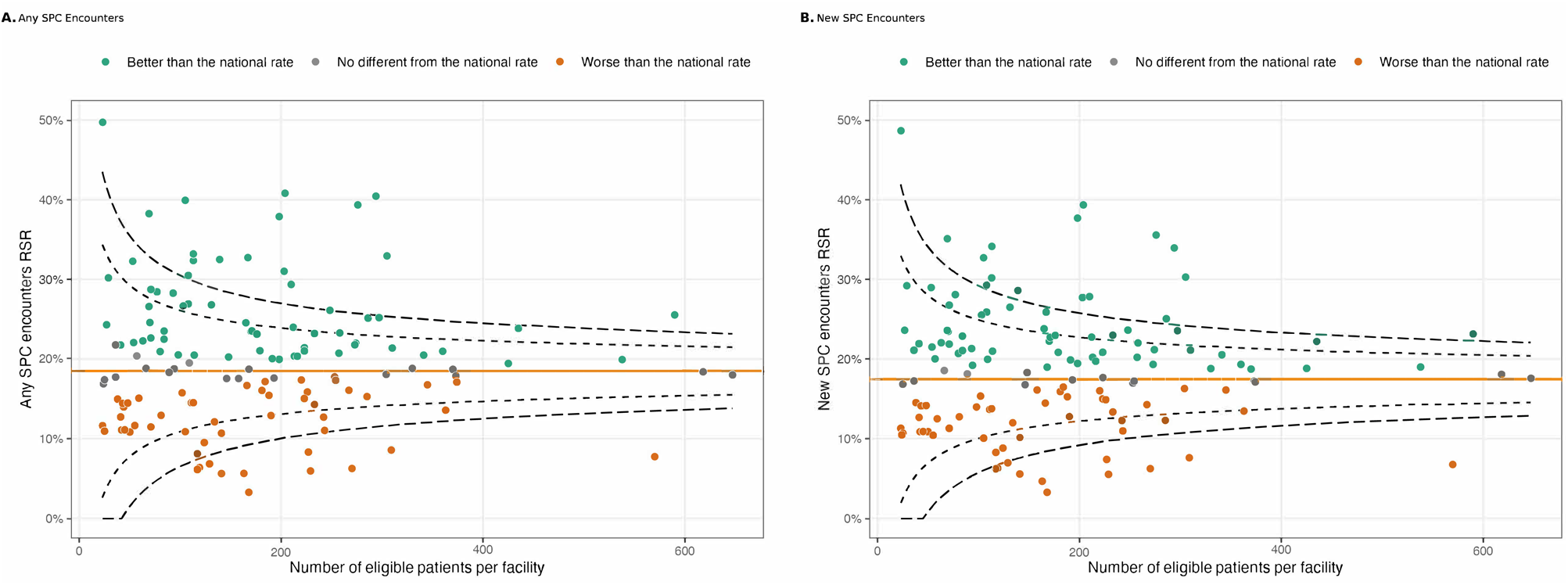
Funnel Plots of RSR of Any and New SPC Encounters by Facility Volume. RSR: Risk standardized rate; SPC = Specialist Palliative Care; Each point represents one VA medical center (n=l33), plotted by facility panel size (x-axis) and risk-standardized palliative care rate (y - axis) from the fully adjusted multilevel model. The horizontal orange line indicates the national average rate (18.5% in Panel A; 17.5% in Panel B). The dashed curves represent control limits around the national mean based on binomial sampling variation (inner: 95%; outer: 99.8%), showing where facility rates would be expected to fall if all facilities were performing at the national mean and observed differences reflected chance alone. Point colors reflect each facility’s classification based on its own 95% bootstrap confidence interval.

### Attributable Risk

Facility-level variation accounted for a net difference of approximately 1,271 SPC encounters over the study period (Figure 3, Panel A). At facilities performing above the national mean, an estimated 791 additional encounters occurred beyond what would be expected if those facilities performed at the national average (17.8% of 4,435 observed encounters). Conversely, at facilities performing below the national mean, an estimated 480 encounters that would have been expected under average performance did not occur (10.8%). Patterns were similar for new SPC encounters: 746 additional encounters (17.8% of 4,190 observed) occurred at above-average facilities, while 449 expected encounters (10.7%) were not delivered at below-average facilities. Ten VAMCs accounted for 48.6% of all excess SPC encounters and 45.0% of all excess new SPC encounters. Similarly, the 10 VAMCs with the largest deficits accounted for 52.8–53.8% of encounters not delivered across outcomes.

**Figure 3.**
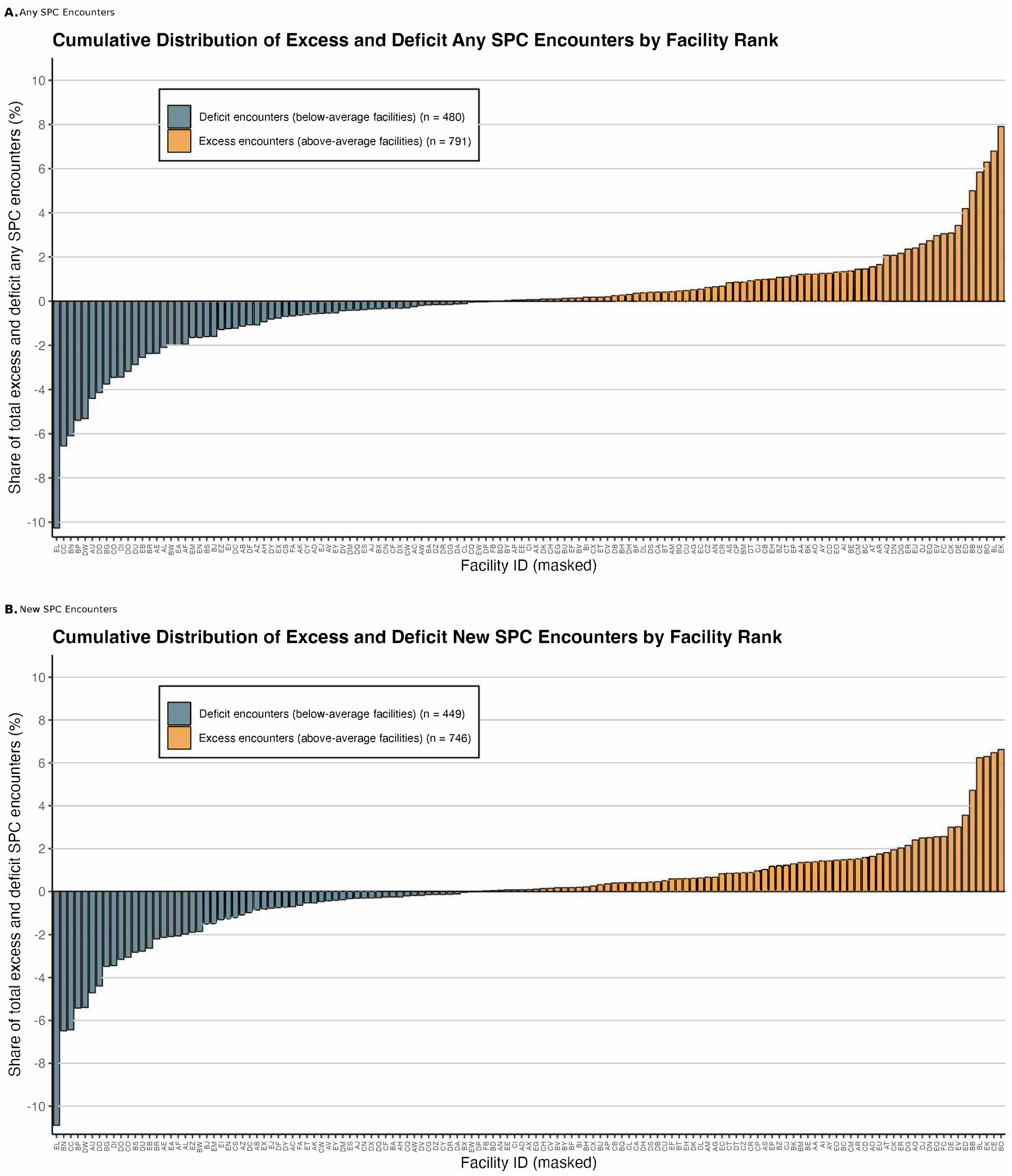
Excess and Deficit Any and New SPC Encounters by Facility. SPC = Specialist Palliative Care; Bidirectional waterfall plots showing each facility’s contribution to deficit (blue, below zero) and excess (orange, above zero) SPC encounters, expressed as a percentage of total deficit or excess encounters nationally. Panel A shows any SPC encounters (480 deficit encounters from below-average facilities, 791 excess encounters from above-average facilities); Panel B shows new SPC encounters (449 deficit, 746 excess). Each bar represents one VA medical center (n=l33). Deficit encounters represent the number of additional encounters that would have occurred if below-average facilities had performed at the national average rate. Excess encounters represent the number of encounters attributable to above-average facility performance beyond what would be expected at the national average rate.

### Sensitivity Analyses

Facility performance classifications were stable across models: 74% (any SPC) and 77% (new SPC) of facilities received the same classification under a model comprised of only patient covariates. In a separate sensitivity analysis comparing each facility against its complexity-tier peers rather than the national mean, the overall share of encounters attributable to above-peer performance decreased to 8.2% for any SPC and 8.9% for new SPC, while the overall share representing below-peer deficits increased to 28.8% and 28.6%, respectively. In a leave-one-out analysis, excluding the largest VAMC from each tier did not substantially change the deficit proportion, supporting the stability of the tier-specific reference despite the small number of facilities per tier (n = 23–26).

## Discussion

Providing palliative care, a guideline-concordant intervention, to patients with aHF is a persistent challenge. Similar to past reports, we found VAMC national average rates of SPC delivery at 17.5%.^4–7^ However, to our knowledge, this is the first study to quantify between-facility variation in SPC delivery for aHF and to estimate the share attributable to measured patient and structural factors. We found that risk-adjusted predicted rates of SPC vary nearly 14-fold across VAMCs, with substantial differences in the odds of receiving palliative care for otherwise similar patients depending on which facility they attend. Most of the between-facility variance (roughly 80%) remains unexplained by patient case-mix or facility structural characteristics alone. Study findings indicate that where a patient with aHF receives care has an independent influence on their likelihood of receiving palliative care, and that the resulting disparity represents a concrete, potentially modifiable target for quality improvement. Importantly, the classification of facilities as “no different than the national rate” was made relative to the national mean, which itself reflects considerable underuse of SPC for aHF; even facilities performing in line with their peers are likely delivering less SPC than is clinically warranted.

ICCs reflect the proportion of total variation in palliative care receipt attributable to differences between facilities rather than differences between patients. The between-facility ICCs of 11.1% and 11.5% for any and new SPC encounters may appear modest but substantially exceed those reported for most cardiovascular outcomes. For context, hospital-level ICCs for heart failure 30-day mortality and readmission are typically 0–3%, and even process-sensitive outcomes, such as persistent critical illness in VA ICUs, have ICCs of only 6.7%.^18–20^ Translating the facility-level variance to the odds ratio scale provides a more clinically interpretable measure of its magnitude: the aMOR of 1.84 and 1.87 indicates that for two patients with identical case-mix, the one cared for at a higher-performing facility has 84–87% greater odds of receiving palliative care. These MORs exceed those reported for hospice use among patients with metastatic cancer (aMOR 1.48), ^21^ guideline-directed medical therapy prescribing in aHF (aMOR 1.23),^22^ health status achievement in outpatient aHF (aMOR 1.70),^23^ and 30-day mortality after cardiac surgery (aMOR 1.20).^20^ Notably, the facility random effect explained more outcome variance than any individual patient-level covariate in the fully adjusted model, suggesting that the medical center where a patient receives care is an important determinant of palliative care delivery, beyond patient-level demographic or clinical characteristics captured in this study.

Our results also add to the broader literature on palliative care delivery in aHF by showing that underuse is not solely a function of the clinical complexity or prognostic uncertainty inherent to this population. Differences in facility-level processes, organizational culture, or care delivery practices, rather than patient severity or institutional resources, explain a meaningful share of this variation. Although aHF differs from cancer in trajectory, treatment intensity, and timing of decline, the observed spread in aHF risk-standardized rates of SPC suggests that some facilities have already achieved substantially greater integration of SPC into usual care. This demonstrates that higher use is feasible within the same national system, under a similar benefit structure and policy context. Accordingly, the gap between high- and low-performing facilities should not be interpreted as inevitable. Rather, variation itself points to an opportunity to make palliative care delivery for HF more consistent, earlier, and more equitable across sites.

Deficits in the receipt of SPC were concentrated among a few facilities: the top 10 facilities with the largest shortfalls accounted for over 50% of all prevented SPC encounters. Lower-performing facilities were distributed across all complexity tiers, arguing against a simple explanation based on size or structural capacity alone. Moreover, in sensitivity analyses, VAMCs were benchmarked against complexity-tier peers rather than the national mean; the proportion of deficit encounters nearly tripled, indicating that most of the palliative care gap arises from within-tier underperformance rather than between-tier structural differences. Together, these findings suggest that targeted interventions at a limited number of sites could yield disproportionate system-wide gains.

Facilities performing below their structural peers represent identifiable, actionable targets for quality improvement. Potential strategies to improve access to SPC may include improved identification of severe disease, better recognition of unmet palliative needs, timely and well-framed referral conversations, favorable cultural local norms regarding palliative care as complementary to disease-directed treatment, and embedding SPC in routine heart failure workflows.^24,25^ The concentration of excess and prevented encounters within a subset of facilities also suggests that positive deviance approaches, methods successfully used in other types of cardiovascular research, may be especially informative.^26,27^ For example, studying centers with consistently high risk-standardized rates could identify scalable practices, staffing models, or referral pathways that are transferable to lower-performing sites.

Several limitations merit consideration. The proportion of facilities classified as performing significantly different from the national mean was high, likely reflecting greater between-facility variance in SPC delivery (ICC = 11.1%) compared with heart failure mortality or readmission (ICC = 0–3%). ^18–20^ Our analysis relied on validated administrative data to identify SPC encounters; however, this approach may not capture all palliative care services delivered. Patients may have received palliative care services outside the VA, episodes that would not have been captured in this analysis – a limitation that applies to the study of any specific healthcare delivery system. Similarly, although we used validated algorithms to capture advanced aHF, misidentification may have occurred. We could not determine the appropriateness of palliative care from administrative data alone, and some variation may reflect appropriate tailoring to patient preferences. Our study examined VA care for veterans, and patterns of variation may differ in other healthcare systems or patient populations. VA’s integrated structure and the availability of universal palliative care make the observed variation particularly striking; variation may be even greater in fragmented healthcare delivery systems.

Facility performance may change over time as staffing, leadership, and organizational priorities evolve. Longitudinal studies of temporal trends in facility performance would provide insights into sustainability and whether variation is increasing or decreasing. Although our case-mix adjustment included several important patient factors, residual confounding remains possible. Finally, our study identified variation and estimated its magnitude, but could not directly examine the mechanisms generating this variation or test specific interventions.

## Conclusion

In this national study of Veterans with aHF, receipt of SPC varied more than 14-fold across VA medical centers. These findings suggest that meaningful differences in healthcare provider behavior, local practice patterns, care processes, and organizational culture help shape whether patients receive guideline-concordant SPC. Because much of the shortfall was concentrated in a relatively small number of facilities, meaningful improvement in SPC delivery in aHF may be feasible through efforts targeted at those relatively lower-performing sites.

## Data Availability

Data are not available at this time.

## Acknowledgements

The authors acknowledge the use of Claude Sonnet 4.6 for code generation and syntax refinement.

## Disclosures

The authors have no conflicts of interest to report.

## Notes

**Sponsor’s Role:** The analysis described here is based on work supported by the National Heart, Lung, and Blood Institute, the Yale Center for Implementation Science, and the Department of Veterans Affairs, Veterans Health Administration, which had no role in the design, methods, participant recruitment, data collection, analysis, or preparation of this article or in the decision to submit this article for publication. The views expressed in this article are those of the authors and do not necessarily reflect the position or policy of the United States Department of Veterans Affairs or the United States Government.

### Competing Interest Statement

The authors have declared no competing interest.

### Author Declarations

The IRBs of Yale University and the West Haven Connecticut, Department of Veterans Affairs gave ethical approval for this work.

## References

1. Virani SS, Alonso A, Benjamin EJ, et al; American Heart Association Council on Epidemiology and Prevention Statistics Committee and Stroke Statistics Subcommittee. Heart disease and stroke statistics—2020 update: a report from the American Heart Association. Circulation. 2020;141(9):e139–e596. Doi: 10.1161/CIR.0000000000000757

2. Sayed A, Abramov D, Fonarow GC, Mamas MA, Kobo O, Butler J, Fudim M. Reversals in the decline of heart failure mortality in the US, 1999 to 2021. JAMA Cardiol. 2024;9(6):585–589. Doi: 10.1001/jamacardio.2024.0615

3. Siddiqi TJ, Minhas AMK, Greene SJ, et al. Trends in heart failure–related mortality among older adults in the United States from 1999–2019. JACC Heart Fail. 2022;10(11):851–859. Doi: 10.1016/j.jchf.2022.06.012

4. Warraich HJ, Godfrey S, Makwana B, et al. Association of palliative care consultation in patients with heart failure with preserved ejection fraction with symptom burden and health care use. JACC Adv. 2025;4(1):101431. Doi: 10.1016/j.jacadv.2024.101431

5. Murthy N, Sedhom R, Parwani P, et al. A nationwide retrospective analysis of trends in palliative care consultation and do-not-resuscitate status in heart failure hospitalizations. Palliat Med. Published online February 21, 2026. Doi: 10.1177/02692163261416339

6. Heidenreich PA, Bozkurt B, Aguilar D, et al. 2022 AHA/ACC/HFSA guideline for the management of heart failure. J Card Fail. 2022;28(5):e1–e167. Doi: 10.1016/j.cardfail.2022.02.010

7. Feder SL, Murphy TE, Abel EA, Akgün KM, Warraich HJ, Ersek M, Fried T, Redeker NS. Incidence and trends in the use of palliative care among patients with reduced, middle-range, and preserved ejection fraction heart failure. J Palliat Med. 2022;25(12):1774–1781. Doi: 10.1089/jpm.2022.0093

8. Feder SL, Han L, Zhan Y, et al. Variation in specialist palliative care reach and associated factors among people with advanced heart failure in the Department of Veterans Affairs. J Pain Symptom Manage. 2024;68(1):22–31.e1. doi: 10.1016/j.jpainsymman.2024.03.022

9. Alqahtani F, Balla S, Almustafa A, Sokos G, Alkhouli M. Utilization of palliative care in patients hospitalized with heart failure: a contemporary national perspective. Clin Cardiol. 2019;42(1):136–142. Doi: 10.1002/clc.23119

10. Sullivan DR, Teno JM, Reinke LF. Evolution of palliative care in the Department of Veterans Affairs: lessons from an integrated health care model. J Palliat Med. 2022;25(1):15–20. Doi: 10.1089/jpm.2021.0246

11. Benchimol EI, Smeeth L, Guttmann A, et al; RECORD Working Committee. The Reporting of studies Conducted using Observational Routinely-collected Health Data (RECORD) statement. PloS Med. 2015;12(10):e1001885. Doi: 10.1371/journal.pmed.1001885

12. Dunlay SM, Blecker S, Schulte PJ, Redfield MM, Ngufor CG, Glasgow A. Identifying patients with advanced heart failure using administrative data. Mayo Clin Proc Innov Qual Outcomes. 2022;6(2):148–155. Doi: 10.1016/j.mayocpiqo.2022.02.001

13. Feder SL, Zhan Y, Abel EA, Smith D, Ersek M, Fried T, Redeker NS, Akgün KM. Validation of electronic health record-based algorithms to identify specialist palliative care within the Department of Veterans Affairs. J Pain Symptom Manage. 2023;66(4):e475–e483. Doi: 10.1016/j.jpainsymman.2023.06.023

14. Osborne TF, Veigulis ZP, Ware A, Arreola DM, Curtin C, Yeung M. Automated electronic health record score to predict mortality risk at the US Department of Veterans Affairs. Am J Hosp Palliat Care. 2025;42(3):230–235. Doi: 10.1177/10499091241247841

15. Kaufman BG, Woolson S, Stanwyck C, et al. Veterans’ use of inpatient and outpatient palliative care: the national landscape. J Am Geriatr Soc. 2024;72(11):3385–3397. Doi: 10.1111/jgs.19141

16. Aldridge MD, Bradley EH. Epidemiology and patterns of care at the end of life: rising complexity, shifts in care patterns and sites of death. Health Aff (Millwood). 2017;36(7):1175–1183. Doi: 10.1377/hlthaff.2017.0182

17. Keenan PS, Normand SL, Lin Z, et al. An administrative claims measure suitable for profiling hospital performance on the basis of 30-day all-cause readmission rates among patients with heart failure. Circ Cardiovasc Qual Outcomes. 2008;1(1):29–37. Doi: 10.1161/CIRCOUTCOMES.108.802686

18. Korda RJ, Du W, Day C, Page K, Macdonald PS, Banks E. Variation in readmission and mortality following 18ospitalization with a diagnosis of heart failure: prospective cohort study using linked data. BMC Health Serv Res. 2017;17(1):220. Doi: 10.1186/s12913-017-2152-0

19. Viglianti EM, Bagshaw SM, Bellomo R, et al. Hospital-level variation in the development of persistent critical illness. Intensive Care Med. 2020;46(8):1567–1575. Doi: 10.1007/s00134-020-06129-9

20. Sanagou M, Wolfe R, Forbes A, Reid CM. Hospital-level associations with 30-day patient mortality after cardiac surgery: a tutorial on the application and interpretation of marginal and multilevel logistic regression. BMC Med Res Methodol. 2012;12:28. Doi: 10.1186/1471-2288-12-28

21. Hua M, Guo L, Ing C, Wang S, Morrison RS. Variation in palliative care program performance for patients with metastatic cancer. J Pain Symptom Manage. 2025;69(1):23–33.e2. doi: 10.1016/j.jpainsymman.2024.10.021

22. Polsinelli VB, Sun JL, Greene SJ, et al. Hospital heart failure medical therapy score and associated clinical outcomes and costs. JAMA Cardiol. 2024;9(11):1029–1038. Doi: 10.1001/jamacardio.2024.2969

23. Khariton Y, Hernandez AF, Fonarow GC, et al. Health status variation across practices in outpatients with heart failure: insights from the CHAMP-HF (Change the Management of Patients With Heart Failure) Registry. Circ Cardiovasc Qual Outcomes. 2018;11(4):e004668. Doi: 10.1161/CIRCOUTCOMES.118.004668

24. Feder SL, Iannone L, Lendvai D, et al. Clinician insights into effective components, delivery characteristics, and implementation strategies of ambulatory palliative care for people with heart failure: a qualitative analysis. J Card Fail. 2025;31(4):611–620. Doi: 10.1016/j.cardfail.2024.07.009

25. Chuzi S, Pensa AV, Allen LA, Cross SH, Feder SL, Warraich HJ. Palliative care for patients with heart failure: results from a Heart Failure Society of America survey. J Card Fail. 2023;29(1):112–115. Doi: 10.1016/j.cardfail.2022.06.010

26. Kassie AM, Eakin E, Abate BB, et al. The use of positive deviance approach to improve health service delivery and quality of care: a scoping review. BMC Health Serv Res. 2024;24(1):438. Doi: 10.1186/s12913-024-10850-2

27. Bradley EH, Curry LA, Ramanadhan S, Rowe L, Nembhard IM, Krumholz HM. Research in action: using positive deviance to improve quality of health care. Implement Sci. 2009;4:25. Doi: 10.1186/1748-5908-4-25

